# Missense variants in *PLA2G6* contribute to a spectrum of clinical syndromes and provide pharmacogenomic correlates

**DOI:** 10.1101/2023.04.06.23288108

**Authors:** Meghana Janardhanan, Padmanabhan Anbhazagan, Biju Viswanath, Padmanabhan Balasundaram, Sanjeev Jain, Meera Purushottam

**Affiliations:** Molecular Genetics Laboratory, Department of Psychiatry, National Institute of Mental Health and Neurosciences, Bangalore, India; Experimental Drug Development Centre, Singapore; Department of Biophysics, National Institute of Mental Health and Neurosciences, Bangalore, India

## Abstract

**Background:** Risk alleles in a gene for a genetic disorder can often cause a spectrum of syndromes. The number of copies, deleteriousness and position in the sequence could influence phenotype manifestation.

**Methods:** Whole exome sequencing in 310 individuals from 100 families with severe mental illness revealed 851 instances of variants in the *PLA2G6* gene. We assessed the population frequency and deleteriousness of the nonsynonymous variants using *in-silico* prediction methods. Molecular docking analyses with antipsychotics was performed to investigate possible pharmacogenomic implications of the *PLA2G6* mutations identified.

**Results:** We found six nonsynonymous variants predicted to be deleterious by VarSome. The frequency of non-synonymous variants was found to vary across populations. The preliminary molecular docking analysis suggests that chlorpromazine and risperidone are predicted to bind at three drug-binding sites however, risperidone has a greater binding affinity to PLA2G6. The occurrence of variants close to these drug-binding sites suggests a possible mechanism for the mediation of parkinsonian side effects on drug intake in patients harboring these variants.

**Conclusion:** Variants in the *PLA2G6*, a gene previously known to be associated with Parkinson’s disease may thus contribute to the risk of psychiatric phenotypes, as observed in these 9 individuals from 6 families with severe mental illness.

## Introduction

Many risk alleles for genetic diseases seem to produce a spectrum of syndromes, depending on whether they occur as heterozygous, homozygous or compound heterozygotes. This may also be linked to differences in phenotype manifestation and the prevalence of these rare damaging variants. Variants in the *PLA2G6* gene are a good example of this.

The *PLA2G6* protein contains 806 amino acids and harbors various domains including ankyrin repeats, a GXSXG lipase catalytic site, a nucleotide-binding domain, and two binding sites for calmodulin (https://www.uniprot.org/). The gene is highly conserved (dN/dS ratio 2.464; missense variant Z-Score 1.21 (gnomAD)), and is syntenic across many vertebrates. The disease-linked variants may contribute to clinical presentation, by modifying the structure and the function of the enzyme.

Neurodegenerative syndromes related to mutations in the *PLA2G6* gene (homozygous and heterozygous) have been described in humans [OMIM 603604] [Guo et al., 2018] and animals [OMIA 002105-9615, OMIA 002105-9940]. Young-onset INAD, early adult life dystonia-PD complex (Park14) and late-onset parkinsonism have all been associated with these variants, and both homozygous and heterozygous states have been implicated. Abnormalities in metabolic processes, phospholipid biosynthesis and membrane remodeling have been implicated in many models of Parkinson’s disease (PD) and schizophrenia. Experimental models that disrupt mitochondrial function (rotenone treatment), and many mutations that affect lipid pathways (*MAPT/PLA2G6*, synuclein etc.) are also known to predispose to PD syndromes [Mori et al., 2019, Magrinelli et al 2022]. Increased phospholipase A2 (PLA2) activity has been reported in schizophrenia, and treatment with antipsychotic drugs reduces the enzyme activity to levels similar to those in unaffected individuals [Gattaz et al., 1987]. Certain networks in the brain may be sensitive to metabolic perturbations, leading to a propensity to develop clinical symptoms.

We assessed the frequency and deleteriousness of variants using *in-silico* prediction methods in the PLA2G6 gene in a sample of patients and controls. Molecular docking analyses with antipsychotics was performed to investigate possible pharmacogenomic implications of the *PLA2G6* mutations identified.

## Methods

We examined the Whole Exome Sequencing (WES) data of 310 individuals with severe mental illness (SMI) (cases(n=190); and controls (familial n=60; population n=60)) [Ganesh et al., 2019]. We used *in-silico* prediction tools such as SIFT, LRT, MutationTaster, FATHMM, MetaSVM from VarSome (Kopanos et al., 2019), which utilise sequence homology, evolutionary conservation, physicochemical and structural properties of amino acids, to predict the effect of the variant on protein function, stability and possible pathogenicity. Network analysis of the gene was performed using GeneMANIA, a website that provides information about interacting protein partners and co-expression, pathways involved and possible function. Further, we carried out molecular docking analysis of antipsychotic drugs with the predicted AlphaFold structure of the protein. Ascertaining the binding pockets would help us to make predictions on the effect of the variants if any on drug interactions.

The full-length predicted protein structure of human 85/88 kDa calcium-independent phospholipase A2 was downloaded from https://alphafold.ebi.ac.uk/entry/O60733. The structure was then imported to the Schrodinger Maestro software package (Maestro, Schrödinger, LLC) and the protein was prepared by adding hydrogen atoms and assigning proper bond orders. Prime (Jacobson et al., 2004) was used to fill in missing side chains, and Epik (Shelley et al., 2007) was used to generate protonation states with a pH of 7.0 +/- 2.0. The protein structure was further optimized by PRCG (Polak-Ribier Conjugate Gradient) minimization (Polak and Ribiere, 1969) method with a maximum of 2500 iterations and converge threshold of 0.05. The total energy of the system after minimization was - 168465.094 KJ/mol. SAVES v6.0 server (https://saves.mbi.ucla.edu/) was used to assess the quality of the model. Results from the Ramachandran plot showed 90.1% and 9.1% of the residues to be in the most favoured regions, and additional allowed regions respectively, suggesting that the quality of the model was reliable.

The possible drug-binding sites in the AlphaFold predicted human PLAG26 protein structure were identified using the SiteMap method (Halgren, 2009). Three potential binding sites (site1, site2 and site3) with high druggability scores (DScore >1) have been identified in the catalytic domain of the enzyme, including the interface region. Prior to docking, all the hydrogen atoms were removed from the protein and only the polar hydrogens were added and Gasteiger Charges were computed. The grid box was centered in the catalytic region of the receptor region (X: -15.188 Y: 7.689 Z: 17.443). The number of grid points in XYZ directions was set to 56*40*40 with a grid spacing of 0.909 Å, such that the grid box covered the whole catalytic site as well as the interface region between the catalytic and ankyrin repeat domains. The docking calculations were performed using AutoDockTools (ADT) v1.5.6 (Morris, G., et al., 2009) and AutoDock Vina (Trott, O et al., 2010) with default settings.

## Results

WES in 310 individuals from 100 families with SMI revealed 851 instances (720 intronic, 57 exonic, 56 5’UTR, 18 3’UTR) of variants in the *PLA2G6* gene. Ten exonic variants (4 synonymous and 6 non-synonymous) were seen in 57 individuals. The six non-synonymous variants, their pathogenicity and prevalence were obtained from Varsome and gnomAD respectively (Table 1, 2 and 3).

**Table 1:** The non-synonymous variants, their pathogenicity and prevalence were obtained from Varsome and gnomAD respectively

| No. | Sample | GRCh37_Start | End | Ref | Alt | AAChange.refGene | Zygosity | Diagnosis |
| --- | --- | --- | --- | --- | --- | --- | --- | --- |
| 1 | P1, P2 | chr22:38508565 | 38508565 | C | T | PLA2G6:NM_003560:<br>exon16:c.2222G>A:R741Q | Homozygous | AREP |
| 2 | P3 | chr22:38508566 | 38508566 | G | A | PLA2G6:NM_003560:<br>exon16:c.2221C>T:p.R741W | Homozygous | INAD |
| 3 | P4 | chr22:38541520 | 38541520 | T | C | PLA2G6:NM_003560:<br>exon3:c.A350G:p.H117R | Heterozygous | Schizophrenia |
| 4 | P5 | chr22:38541464 | 38541464 | G | A | PLA2G6:NM_003560:<br>exon3:c.C406T:p.H136Y | Heterozygous | Schizophrenia |
| 5 | C1 | chr22:38536020 | 38536020 | T | C | PLA2G6:NM_003560:<br>exon5:c.A766G:p.I256V | Heterozygous | Control |
| 6 | P6, P7,<br>P8, P9,<br>C2, C3,<br>C4, C5 | chr22:38528888 | 38528888 | C | T | PLA2G6:NM_003560:<br>exon7:c.G1027A:p.A343T | Heterozygous | P6, P7, P8,<br>P9- BPAD;<br>C2,C3, C4,<br>C5- Controls |
| 7 | C6 | chr22:38525511 | 38525511 | G | A | PLA2G6:NM_003560:<br>exon8:c.C1136T:p.P379L | Heterozygous | Control |
| 8 | P10,<br>P11,<br>P12 | chr22:38525518 | 38525518 | C | A | PLA2G6:NM_003560:<br>exon8:c.G1129T:p.D377Y | Heterozygous | Parkinsonism,<br>Schizophrenia |

A novel heterozygous missense mutation p.D377Y (chr22:38525518 C>A) located in the ankyrin repeats region was seen in three ill siblings (P10, P11 and P12) from a family with complex neuropsychiatric symptoms. P10 was diagnosed to have schizophrenia, while P11 had Parkinsonian features with extrapyramidal symptoms and was suicidal, and P12 had epilepsy and psychosis and also exhibited Parkinsonian symptoms towards the end of life. Variants p.H117R and p.H136Y, located in the region preceding the ankyrin repeats, were identified in individuals diagnosed with schizophrenia (P4 and P5). A p.A343T variant located within the ankyrin repeat domain was detected in four individuals diagnosed with the bipolar affective disorder (P6, P7, P8, P9); and also in our healthy young controls (C2, C3, C4, C5). P6, P7 and C2 were all individuals from the same family, Two more variants-p.I256V and p.P379L both located in the ankyrin repeat domain were identified in two different young healthy individuals (C1 and C6), who had no symptoms at the time of assessment.

GeneMania detected a network of twenty genes involved in oxidative stress, mitochondrial dysfunction and lipid remodeling, to be interacting with *PLA2G6*.

The preliminary molecular docking analysis suggests that while both chlorpromazine and risperidone are predicted to bind at three drug-binding sites, risperidone has a greater binding affinity (Figure 1). In addition, the positions Arg741 (near site 2) and Asp377 (near site 3) which is the interface region between the catalytic domain and the ankyrin repeat, are near these predicted binding sites. Amino acid changes at these positions as observed in our samples could impact drug binding and biological function. Since the protein probably functions as a dimer (Malley et al, 2018), the effect of these variants may be further amplified. Previously, mutations in this gene have been associated with PD. It is thus plausible that these variants that affected the shape and hence possible protein interactions have triggered parkinsonian symptoms in our patients.

**Figure 1:**
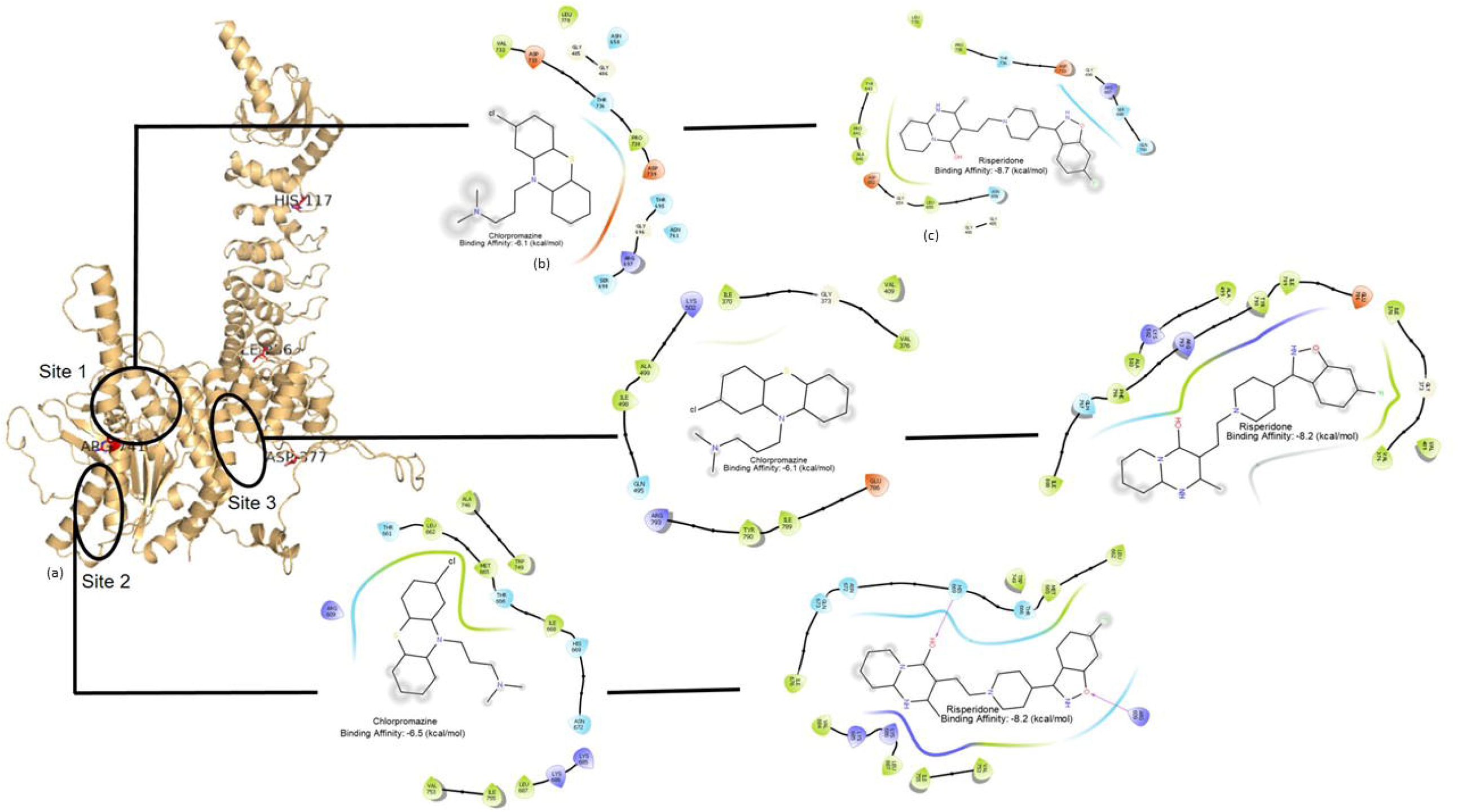
Molecular docking analysis of antipsychotics with the predicted structure of PLA2G6 (a) The final full-length structure is shown with the putative binding sites circled in black; site 3 is the interface region between the catalytic domain and the ankyrin repeat domain. The residues that undergo mutation are labelled. Interactions of chlorpromazine(b) and risperidone (c) in the three different binding sites with the PLA2G6 receptor are shown. Hydrophobic interactions (in b,c) are in green, polar interactions in sky blue, negatively and positively charged residue interactions in red and blue respectively. Docking results show that risperidone has a better binding affinity than chlorpromazine.

## Discussion

Eighteen individuals out of 310 were identified to carry deleterious non-synonymous mutations in the *PLA2G6* gene. Twelve had a diagnosis of a neuropsychiatric condition, while 6 were unaffected (one of whom was related to an ill individual, while the other five were unrelated, healthy controls). Of the 8 variants reported here, p.H136Y, I256V and p.A343T are reported in the GenomeAsia (GenomeAsia100K Consortium, 2019). The p.H136Y and I256V are found only in the South Asian (SAS) population at a low frequency (0.00069 and 0.00003375 respectively) (Supplementary). The p.A343T variant seen in multiple cases and controls has a low prevalence across several populations in gnomAD. The p.D377Y is not reported on Clinvar/gnomAD. We had earlier observed the homozygous p.R741Q variants in two individuals (P1, P2) with AREP [Sakhardande et al., 2021], and a p.R741W variant in an individual (P3) with INAD. These variants lie in the region succeeding the calmodulin-binding domain. It has been proposed that variants in this gene cause a spectrum of phenotypes, as these variants may influence different functions of the protein (Erro et al, 2016; Magrinelli et al 2022).

The Ca2+-independent phospholipase A2 (iPLA2), the enzyme encoded by *PLA2G6* is known to mediate the release of free fatty acids and lysophospholipids by catalyzing the hydrolysis of the sn-2 fatty acyl bond of phospholipids. This plays an important role in phospholipid remodeling, signal transduction, cell proliferation, endoplasmic reticulum stress-mediated apoptosis, and ferroptosis (Wan et al, 2022), by its impact on membrane permeability and fluidity, and ion homeostasis.

The *in-silico* analysis of the pathogenic/deleterious variants of the 8 non-synonymous variants identified in the *PLA2G6* gene shows that these mutations may impact protein structure and function. All eight mutations involving conserved positions were predicted to be damaging and affected protein stability to varying degrees. This would also have an indirect effect on other proteins that interact with it.

The pathobiology of both Parkinson’s disease and schizophrenia is thought to be related to neurotransmitter imbalance at the synapse, but the impact of metabolic processes that interact with synapse function may also be a critical component. Antipsychotic drugs bind membrane receptors, are invaginated, endocytosed and then recycled. Many actions are dependent on this transition through the post-synaptic cellular milieu. Metabolic processes that impact lipid membrane function, and thus endocytic and vacuole formation, would influence sensitivity to antipsychotics. The Parkinsonian side effects occur in a large proportion of patients exposed to antipsychotic drugs and may be influenced by variations in the *PLA2G6* gene.

The two individuals with AREP developed severe Parkinsonian side effects when treated with antipsychotics. This sensitivity to antipsychotic drugs was perhaps linked to the presence of variants in a critical region of the protein that binds to these drugs (and perhaps to the endogenous ligand, dopamine). Dopamine-mediated regulation of intracellular lipid dynamics may thus be an important factor in integrating signaling and metabolic function. Long-term dysfunction of this process, by homozygous variants, may underlie the progressive nature of the AREP/Park14 syndrome. The extreme sensitivity to antipsychotics may be one pharmacogenomic aspect of this variation. Other variants (less damaging but more common) may even underlie the differences in sensitivity to parkinsonian side effects in the general population.

Variants in the PLA2G6 gene may thus contribute to the risk of psychiatric phenotypes, as observed in these 9 individuals from 6 families. Many of these individuals showed parkinsonian symptoms during follow-up. This seems to suggest that the continuum of phenotypes described for this gene might even extend to drug-induced side effects. We suggest that variants in the PLA2G6 gene be investigated with reference to the propensity to develop parkinsonian side effects on treatment with antipsychotics.

Thus, long-term follow-up is essential to understand the effect of rare, damaging variants over a lifetime. Reports also suggest the enrichment of particular gene variants in different populations (Chu et al, 2020). This is interesting and might need an extensive compilation of mutation databases to understand the differences in clinical presentation and epidemiology of these genetic variants across the world. Modelling the links between genetic variation, protein structure, and the impact on cell biology can thus help understand the risk of disease, its progression, drug response, and side effects.

## Data Availability

The datasets generated and/or analysed during the current study are available from the ADBS bio repository https://www.ncbs.res.in/adbs/bio-repository on registration and reasonable request.

## Author contributions

Conception and Design: MP, SJ and BV; analysis and interpretation of the data: MJ, PA, BP; writing-original draft: MJ; writing-review and editing: MP, SJ, BV

## Acknowledgements

We wish to acknowledge Dr Ravi Yadav, Dr Venkata Senthil Kumar Reddi and Dr Gautham Arunachal for the clinical identification of the cases. The work was supported by the “Accelerator Program for Discovery in Brain Disorders Using Stem Cells”, Department of Biotechnology, Government of India (Grant BT/PR17316/MED/31/326/2015). BV is also funded by a DBT/Wellcome Trust India Alliance Intermediate (Clinical and Public Health) Research Fellowship (IA/CPHI/20/1/505266).

## Notes

### Competing Interest Statement

The authors have declared no competing interest.

### Author Declarations

The study protocol was approved by the Institutional Ethics Committee of National Institute of Mental Health and Neurosciences, Bengaluru, India, and the study procedures conformed to the provisions of the Declaration of Helsinki. All participants provided written informed consent.

