## Supplemental File for "Missense variants in *PLA2G6* contribute to a spectrum of clinical syndromes and provide pharmacogenomic correlates"

**Supplementary Material:**

| No. | Missense Mutation |  | Allele Frequency (gnomAD- accessed on 10/14/2022) | | | | | | | | |
| --- | --- | --- | --- | --- | --- | --- | --- | --- | --- | --- | --- |
|  |  | rsID | Globalallele  frequency | South Asian | East Asian | European (Finnish) | European (non-Finnish) | African/ African American | Latino/ Admixed American | Ashkenazi Jewish | Others |
| 1 | p.R741Q | rs121908686 | 0.00009033 | 0.0005175 | 0 | 0 | 3.37E-05 | 0 | 0 | 0 | 0 |
| 2 | p.R741W | rs530348521 | 0.000006465 | 0 | 0 | 0 | 0 | 0 | 3.96E-05 | 0 | 0 |
| 3 | p.H117R | rs750384765 | 0.00002822 | 0.000199 | 0 | 0 | 0 | 0 | 2.93E-05 | 0 | 0 |
| 4 | p.H136Y | rs566979554 | 0.0000453 | 0.0003719 | 0 | 0 | 0 | 0 | 0 | 0 | 0 |
| 5 | p.I256V | rs777270576 | 0.00000415 | 0.00003375 | 0 | 0 | 0 | 0 | 0 | 0 | 0 |
| 6 | p.A343T | rs11570680 | 0.01056 | 0.01516 | 0.0001493 | 0.007798 | 0.01388 | 0.002197 | 0.005772 | 0.01977 | 0.01288 |
| 7 | p.P379L | rs570834054 | 0.00002386 | 0.000196 | 0 | 0 | 0 | 0 | 0 | 0 | 0 |
| 8 | p.D377Y | . | . | . | . | . | . | . | . | . | . |

Table 1: The prevalence of the 8 non-synonymous variants from gnomAD.

| **No.** | **Missense Mutation** | **Pathogenicity Prediction** | | | |
| --- | --- | --- | --- | --- | --- |
|  |  | **SIFT** | **MutationTaster** | **FATHMM** | **MetaSVM** |
| 1 | p.R741Q | T | T | Uncertain | Uncertain |
| 2 | p.R741W | Uncertain | T | Uncertain | Uncertain |
| 3 | p.H117R | D | D | T | T |
| 4 | p.H136Y | T | D | T | T |
| 5 | p.I256V | D | D | T | T |
| 6 | p.A343T | T | D | T | T |
| 7 | p.P379L | T | D | T | T |
| 8 | p.D377Y | D | D | T | T |

Table 2: The pathogenicity predictions obtained from Varsome. D- deleterious, T- tolerated

References for the tools and databases employed:

| Tool/Database | Reference |
| --- | --- |
| gnomAD | Karczewski, K.J., Francioli, L.C., Tiao, G. et al. The mutational constraint spectrum quantified from variation in 141,456 humans. Nature 581, 434–443 (2020). https://doi.org/10.1038/s41586-020-2308-7 |
| VarSome | Kopanos, C., Tsiolkas, V., Kouris, A., Chapple, C. E., Albarca Aguilera, M., Meyer, R., & Massouras, A. (2019). VarSome: the human genomic variant search engine. Bioinformatics (Oxford, England), 35(11), 1978–1980. https://doi.org/10.1093/bioinformatics/bty897 |
| PolyPhen2 | Adzhubei IA, Schmidt S, Peshkin L, et al. A method and server for predicting damaging missense mutations. Nat Methods. 2010;7(4):248-249. doi:10.1038/NMETH0410-248 |
| Align GVGD | Mathe, E., Olivier, M., Kato, S., Ishioka, C., Hainaut, P., & Tavtigian, S. V. (2006). Computational approaches for predicting the biological effect of p53 missense mutations: a comparison of three sequence analysis based methods. Nucleic acids research, 34(5), 1317–1325.<https://doi.org/10.1093/nar/gkj518> |
| MUpro | Cheng J, Randall A, Baldi P. Prediction of protein stability changes for single-site mutations using support vector machines. Proteins Struct Funct Genet. 2006;62(4):1125-1132. doi:10.1002/PROT.20810 |
| I-Mutant Suite | Capriotti E, Fariselli P, Casadio R. I-Mutant2.0: Predicting stability changes upon mutation from the protein sequence or structure. Nucleic Acids Res. 2005;33(SUPPL. 2). doi:10.1093/NAR/GKI375 |
| GeneMania | Warde-Farley D, Donaldson SL, Comes O, et al. The GeneMANIA prediction server: Biological network integration for gene prioritization and predicting gene function. Nucleic Acids Res. 2010;38(SUPPL. 2). doi:10.1093/NAR/GKQ537 |
| Epik | Shelley JC, Cholleti A, Frye LL, Greenwood JR, Timlin MR, Uchimaya M J Comput Aided Mol Des. 2007 Dec; 21(12):681-91. |
| PRCG (Polak-Ribier Conjugate Gradient) minimization | E. Polak & G. Ribiere, Revenue Francaise Informat. Recherche Operationelle, 16, 35 (1969) |
| SiteMap Method | Halgren TA  J Chem Inf Model. 2009 Feb; 49(2):377-89. |
